# Changes in child and adolescent mental health across the COVID-19 pandemic (2018-2023): Insights from general population and clinical samples in the Netherlands

**DOI:** 10.1101/2023.08.29.23294764

**Authors:** Hedy A. van Oers, Hekmat Alrouh, Jacintha M. Tieskens, Michiel A.J. Luijten, Rowdy de Groot, Emma Broek, Daniël van der Doelen, Helen Klip, Ronald De Meyer, Malindi van der Mheen, I. Hyun Ruisch, Germie van den Berg, Hilgo Bruining, Jan Buitelaar, Rachel van der Rijken, Pieter J. Hoekstra, Marloes Kleinjan, Ramón Lindauer, Kim J. Oostrom, Wouter Staal, Robert Vermeiren, Ronald Cornet, Lotte Haverman, Arne Popma, Meike Bartels, Tinca J. C. Polderman, Josjan Zijlmans

## Abstract

**Background:** The COVID-19 pandemic negatively affected child and adolescent mental health and at the end of the pandemic (April 2022) child mental health had not returned to pre-pandemic levels. We investigated whether this observed increase in mental health problems has continued, halted, or reversed after the end of the pandemic in children from the general population and in children in psychiatric care.

**Methods:** We collected parent-reported and child-reported data at two additional post-pandemic time points (November/December 2022 and March/April 2023) in children (8-18 years) from two general population samples (N=818-1056 per measurement) and one clinical sample receiving psychiatric care (N=320-370) and compared these with data from before the pandemic. We collected parent-reported data on internalizing and externalizing problems with the Brief Problem Monitor (BPM) and self-reported data on Anxiety, Depressive symptoms, Sleep-related impairments, Anger, Global health, and Peer relations with the Patient-Reported Outcomes Measurement Information System (PROMIS®).

**Results:** In the general population, parents reported no changes in externalizing problems but did report higher internalizing problems post-pandemic than pre-pandemic. Children also reported increased mental health problems post-pandemic, especially in anxiety and depression, to a lesser extent in sleep-related impairment and global health, and least in anger. In the clinical sample, parents reported higher internalizing, but not externalizing problems post-pandemic compared to the start of the pandemic. Children reported greatest increases in problems in anxiety, depression, and global health, to a lesser extent on sleep-related impairment, and least on anger.

**Conclusions:** Child mental health problems in the general population are substantially higher post-pandemic compared to pre-pandemic measurements. In children in psychiatric care mental health problems have increased during the pandemic and are substantially higher post-pandemic than at the start of the pandemic. Longitudinal and comparative studies are needed to assess what the most important drivers of these changes are.

## Introduction

The COVID-19 pandemic had a severe impact on society. Children and adolescents (hereafter referred to as children) were affected by the pandemic because of restrictions such as lockdowns, school closures, and physical distancing. Given their stage of development, children were shown to be particularly vulnerable to negative mental health effects. In fact, within the first two months after the start of the pandemic, children reported more feelings of anxiety and depression compared to pre-pandemic time points (Luijten, van Muilekom, et al. 2021; Shoshani and Kor 2022). Meta-analyses and reviews conducted over a more extended period of time have shown the persistence of these problems throughout the pandemic (Kauhanen et al. 2023; Harrison et al. 2022; Samji et al. 2022; Deng et al. 2023; Newlove-Delgado et al. 2023; Ludwig-Walz et al. 2023).

Previously, we examined the trajectory of child mental health in three large clinical or general population samples from the start of the COVID-19 pandemic up to two years into it (Zijlmans et al. 2023). Our findings showed that in the general population in particular internalizing mental problems were most prevalent during the initial year, had since started to improve, but had not yet returned to pre-pandemic levels by April 2022. Conversely, in a psychiatric care sample, these problems continued to increase throughout the pandemic. Our general population findings are in line with a recent meta-analysis showing a positive correlation between date of data collection and the prevalence of anxiety and depressive symptoms throughout the pandemic, with a slight downward trend since the winter semester of 2021 (Deng et al. 2023). However, the pattern of mental problems over time in clinical populations are yet unclear due to limited studies in this area.

Here, we aimed to extend our findings and assess whether the observed trend towards pre-pandemic levels of mental health in the general population (Zijlmans et al. 2023), and the observed increase in internalizing mental health problems in children in psychiatric care has continued, halted, or reversed. Therefore, we collected mental health data at two additional post-pandemic time points (November/December 2022 and March/April 2023) in children from the general population and in children receiving psychiatric care and compared these with data from before the COVID-19 pandemic. We collected parent-reported and self-reported outcome measures on multiple domains of mental health, covering internalizing and externalizing problems.

## Materials and methods

The Dutch consortium Child and Adolescent Mental Health and WellBeing in times of the COVID-19 pandemic (CAMHWB-19) studied two general population samples and one clinical sample of (parents of) children aged 8-18 years. We previously reported on the first five pandemic time points of this study, ranging from April 2020 to April 2022 (Zijlmans et al. 2023). More details on the two general population samples, (the Netherlands Twin Register (NTR) and the KLIK group which collected samples using an online panel agency), as well as the clinical sample of children receiving psychiatric care, DREAMS (Dutch REsearch in child and Adolescent Mental health), can be found in this previous report (Zijlmans et al. 2023).

### Participants

Participants were children between 8 and 18 years old, with a mean age across all time points of 11.5 years in the NTR sample, 13.7 years in the KLIK sample, and 13.4 years in the DREAMS sample. For detailed information on the samples, see Table 1. Note that numbers may differ slightly compared to our previous report due to updated datasets. We received approval for data collection from the appropriate ethics committees and all children and parents provided informed consent. The studies were conducted in accordance with the ethical standards outlined in the 1964 Declaration of Helsinki and its later amendments.

**Table 1.**
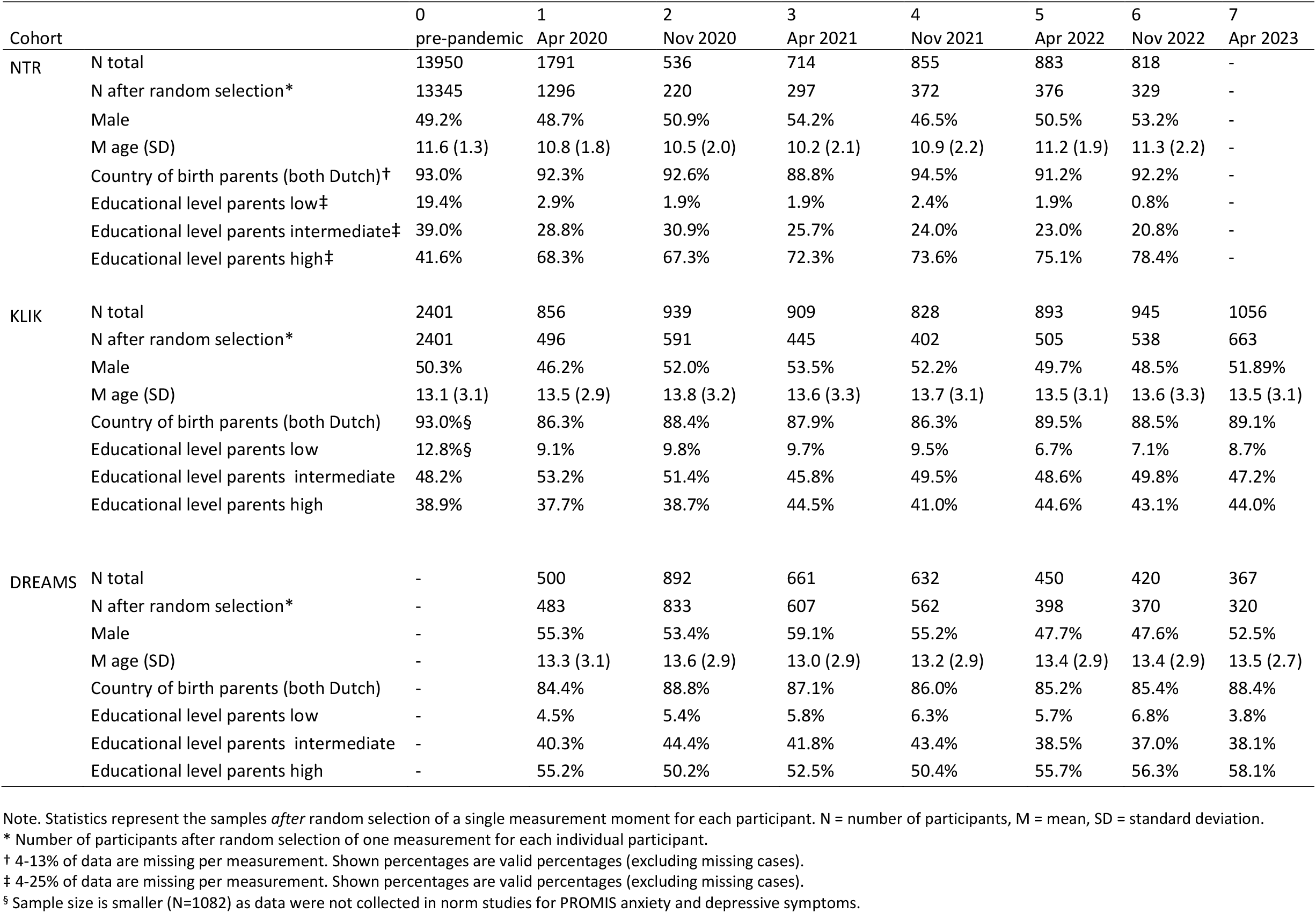
Sociodemographic Characteristics of the Samples.

| Cohort |  | 0<br>pre-pandemic | 1<br>Apr 2020 | 2<br>Nov 2020 | 3<br>Apr 2021 | 4<br>Nov 2021 | 5<br>Apr 2022 | 6<br>Nov 2022 | 7<br>Apr 2023 |
| --- | --- | --- | --- | --- | --- | --- | --- | --- | --- |
| NTR | N total | 13950 | 1791 | 536 | 714 | 855 | 883 | 818 | - |
|  | N after random selection* | 13345 | 1296 | 220 | 297 | 372 | 376 | 329 | - |
|  | Male | 49.2% | 48.7% | 50.9% | 54.2% | 46.5% | 50.5% | 53.2% | - |
|  | M age (SD) | 11.6 (1.3) | 10.8 (1.8) | 10.5 (2.0) | 10.2 (2.1) | 10.9 (2.2) | 11.2 (1.9) | 11.3 (2.2) | - |
|  | Country of birth parents (both Dutch)† | 93.0% | 92.3% | 92.6% | 88.8% | 94.5% | 91.2% | 92.2% | - |
|  | Educational level parents low‡ | 19.4% | 2.9% | 1.9% | 1.9% | 2.4% | 1.9% | 0.8% | - |
|  | Educational level parents intermediate‡ | 39.0% | 28.8% | 30.9% | 25.7% | 24.0% | 23.0% | 20.8% | - |
|  | Educational level parents high‡ | 41.6% | 68.3% | 67.3% | 72.3% | 73.6% | 75.1% | 78.4% | - |
| KLIK | N total | 2401 | 856 | 939 | 909 | 828 | 893 | 945 | 1056 |
|  | N after random selection* | 2401 | 496 | 591 | 445 | 402 | 505 | 538 | 663 |
|  | Male | 50.3% | 46.2% | 52.0% | 53.5% | 52.2% | 49.7% | 48.5% | 51.89% |
|  | M age (SD) | 13.1 (3.1) | 13.5 (2.9) | 13.8 (3.2) | 13.6 (3.3) | 13.7 (3.1) | 13.5 (3.1) | 13.6 (3.3) | 13.5 (3.1) |
|  | Country of birth parents (both Dutch) | 93.0%§ | 86.3% | 88.4% | 87.9% | 86.3% | 89.5% | 88.5% | 89.1% |
|  | Educational level parents low | 12.8%§ | 9.1% | 9.8% | 9.7% | 9.5% | 6.7% | 7.1% | 8.7% |
|  | Educational level parents intermediate | 48.2% | 53.2% | 51.4% | 45.8% | 49.5% | 48.6% | 49.8% | 47.2% |
|  | Educational level parents high | 38.9% | 37.7% | 38.7% | 44.5% | 41.0% | 44.6% | 43.1% | 44.0% |
| DREAMS | N total | - | 500 | 892 | 661 | 632 | 450 | 420 | 367 |
|  | N after random selection* | - | 483 | 833 | 607 | 562 | 398 | 370 | 320 |
|  | Male | - | 55.3% | 53.4% | 59.1% | 55.2% | 47.7% | 47.6% | 52.5% |
|  | M age (SD) | - | 13.3 (3.1) | 13.6 (2.9) | 13.0 (2.9) | 13.2 (2.9) | 13.4 (2.9) | 13.4 (2.9) | 13.5 (2.7) |
|  | Country of birth parents (both Dutch) | - | 84.4% | 88.8% | 87.1% | 86.0% | 85.2% | 85.4% | 88.4% |
|  | Educational level parents low | - | 4.5% | 5.4% | 5.8% | 6.3% | 5.7% | 6.8% | 3.8% |
|  | Educational level parents intermediate | - | 40.3% | 44.4% | 41.8% | 43.4% | 38.5% | 37.0% | 38.1% |
|  | Educational level parents high | - | 55.2% | 50.2% | 52.5% | 50.4% | 55.7% | 56.3% | 58.1% |

Note. Statistics represent the samples *after* random selection of a single measurement moment for each participant. N = number of participants, M = mean, SD = standard deviation.
\* Number of participants after random selection of one measurement for each individual participant.
† 4-13% of data are missing per measurement. Shown percentages are valid percentages (excluding missing cases).
‡ 4-25% of data are missing per measurement. Shown percentages are valid percentages (excluding missing cases).
§ Sample size is smaller (N=1082) as data were not collected in norm studies for PROMIS anxiety and depressive symptoms.

### Procedure

In the current study, we collected data at two post-pandemic time points (November/December 2022 and March/April 2023) after previously collecting data at five time points during the pandemic (between April 2020 and April 2022). At each time point, new and recurrent participants were invited via email to participate. Response rates varied between 14% and 45% for NTR and between 9% and 11% for DREAMS. For the KLIK sample, the sampling procedure was designed to end up with a representative sample and no meaningful response rate can be calculated. To prevent within-subject effects biasing the results, we randomly selected one time point occasion for each participant in all samples, resulting in a repeated cross-sectional design. A mixed design was not possible as not all samples had enough within-subjects data available. Pre-pandemic data were available for the two general population samples, but not for the clinical sample. The NTR sample does not have data on the second post-pandemic time point (April 2023). Table 1 presents an overview of the samples and data that were used for the analyses. Figure 1 provides an overview of the data collection points in relation to Dutch COVID restrictions at the time.

**Figure 1.**
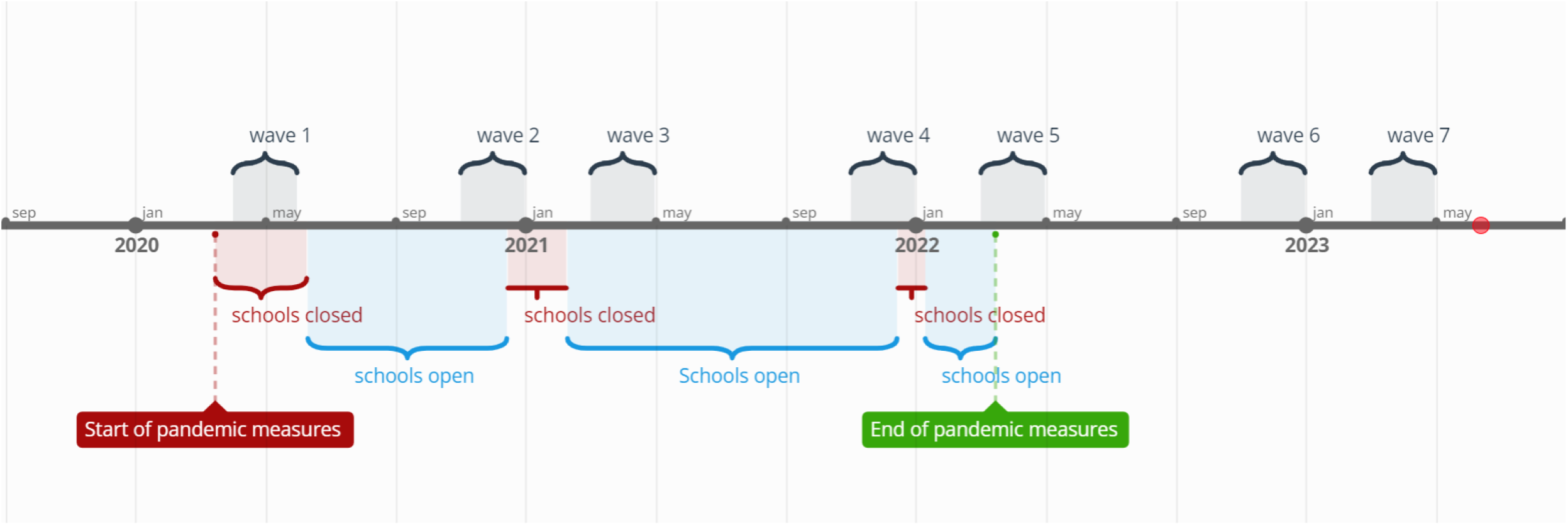

### Measures

#### Socio-demographic information

We collected data on age and sex of the child, and the parent’s country of birth and educational level. Country of birth was defined as both parents being born in the Netherlands (yes/no). Parental educational level was categorized based on the highest education level among both parents and coded as low (primary education, lower vocational education, or lower and middle general secondary education), intermediate (middle vocational education, higher secondary education, or pre-university education), and high (higher vocational education or university).

### Parent-Reported Outcomes

For parental reports in NTR and DREAMS, we employed the Brief Problem Monitor from the Achenbach System of Empirical Based Assessment (ASEBA-BPM). The BPM (Achenbach TM 2011) is a shortened version of the Child Behavior Checklist (CBCL/6-18 years) (Achenbach TM 2001) and assesses behavioral and emotional problems in children as reported by their parents. Items are rated on a three-point Likert-scale, where parents rate if a statement applies to their child (0 = ‘not true’, 1 = ‘somewhat true’, 2 = ‘very true’). In line with the BPM manual, we coded missing items on the BPM as zero. If more than 20% of items were missing for a participant, they were excluded from the BPM analysis. The BPM provides an internalizing score consisting of six items and an externalizing score. The externalizing score typically consists of seven items; however, we excluded one item related to behavior at school due to data collection occurring during periods when children did not attend school. The six remaining items were weighted to maintain the same range as the normal scoring system, allowing for comparison to other studies.

### Child-Reported Outcomes

For child self-reports in the KLIK and DREAMS samples, we employed the Patient-Reported Outcomes Measurement Information System (PROMIS®). Six measures of the Dutch-Flemish PROMIS® were used to assess self-reported mental health: Anxiety v2.0 (Irwin et al. 2010), Depressive Symptoms v2.0 (Irwin et al. 2010), Anger v2.0 (Irwin et al. 2012), Sleep-related impairment v1.0 (Forrest et al. 2018), Global health v1.0 (Forrest et al. 2014), and Peer Relationships v2.0 (Dewalt et al. 2013). All instruments except Anger and Global Health were administered as Computerized Adaptive Tests (CAT), where items are selected based on responses to previously completed items, resulting in reliable scores with fewer items (Cella et al. 2007). Most items are scored on a five-point Likert scale ranging from ‘never’ to ‘(almost) always’. Total scores are calculated by transforming item scores into T-scores ranging from 0 to 100, with a mean of 50 and a standard deviation of 10 in the original calibration sample (Irwin et al. 2010). The U.S. item parameters were used in the CAT algorithm and T-score calculations, as by PROMIS convention. The PROMIS pediatric item banks and scales have previously been validated in the Dutch population (Klaufus et al. 2021; Luijten, van Litsenburg, et al. 2021; van Muilekom et al. 2021; Peersmann et al. 2022; Luijten et al. 2022).

### Data analysis

Statistical analyses were performed using IBM SPSS Statistics 28. Within each sample and for each outcome variable we performed analyses of covariance (ANCOVA) to test for differences in severity and type of mental health problems over the course of the pandemic. In all analyses we included age and sex of the child as covariates and tested for interaction effects between time and sex, as well as time and age. For the latter interaction we categorized age into two groups: 8-12 years and 13-18 years. We performed post-hoc Least Significant Differences tests to compare individual time points within each sample. For the BPM measures differences in scores are reported as estimated marginal means (EMM) of Z-scores standardized to pre-pandemic data of the NTR for ease of interpretation (Table 2). Sum scores are presented in Table S1 to facilitate comparison to other (international) studies. Similarly, differences in scores for the PROMIS measures are reported as EMMs of Z-scores standardized to pre-pandemic norm scores for KLIK (Table 2), with T-scores based on the original (U.S.) calibration sample reported for international comparison in Table S2. Finally, we report the proportions of children who scored outside of the normal range on the BPM internalizing and externalizing scales based on rater and sex specific T-scores (T > 65) in Table S1 and the proportion of children who scored outside of the normal range on the PROMIS scales in Table S3.

**Table 2.** BPM and PROMIS standardized estimated marginal means (EMM) standard errors. and comparisons between measurement points.

| Cohort |  | 0 (a)<br>pre-pandemic | 1 (b)<br>Apr 2020 | 2 (c)<br>Nov 2020 | 3 (d)<br>Apr 2021 | 4 (e)<br>Nov 2021 | 5 (f)<br>Apr 2022 | 6 (g)<br>Nov 2022 | 7 (h)<br>Apr 2023 |
| --- | --- | --- | --- | --- | --- | --- | --- | --- | --- |
| NTR | N | 13345 | 1296 | 220 | 297 | 372 | 376 | 329 | - |
|  | BPM Internalizing | 0 (0.02)<sup>bd-g</sup> | 0.32 (0.04)<sup>ac-g</sup> | 0.09 (0.09)<sup>bd-g</sup> | 0.61 (0.08)<sup>abc</sup> | 0.57 (0.07)<sup>abc</sup> | 0.5 (0.07)<sup>abc</sup> | 0.52 (0.07)<sup>abc</sup> | - |
|  | BPM Externalizing | 0.07 (0.02)<sup>df</sup> | 0.1 (0.04) | -0.07 (0.08)<sup>def</sup> | 0.26 (0.08)<sup>ac</sup> | 0.2 (0.07)<sup>c</sup> | 0.21 (0.07)<sup>ac</sup> | 0.13 (0.07) | - |
| DREAMS | N | - | 431 | 675 | 538 | 488 | 355 | 338 | 294 |
|  | BPM Internalizing | - | 2.84 (0.13)<sup>defh</sup> | 2.96 (0.12)<sup>defh</sup> | 3.27 (0.12)<sup>bc</sup> | 3.31 (0.13)<sup>bc</sup> | 3.3 (0.14)<sup>bc</sup> | 3.1 (0.15)<sup>h</sup> | 3.54 (0.16)<sup>bcg</sup> |
|  | BPM Externalizing | - | 1.32 (0.09) | 1.43 (0.08)<sup>g</sup> | 1.51 (0.09)<sup>g</sup> | 1.48 (0.09)<sup>g</sup> | 1.51 (0.1)<sup>g</sup> | 1.16 (0.11)<sup>c-fh</sup> | 1.47 (0.12)<sup>g</sup> |
| KLIK | N | 527-1082* | 467-482 | 408-423 | 383-387 | 335-345 | 448-455 | 474-485 | 551-577 |
|  | Anxiety<sup>†§</sup> | -0.01 (0.03)<sup>b-h</sup> | 0.66 (0.05)<sup>af</sup> | 0.64 (0.05)<sup>af</sup> | 0.59 (0.05)<sup>a</sup> | 0.70 (0.05)<sup>af</sup> | 0.49 (0.05)<sup>abce</sup> | 0.58 (0.05)<sup>a</sup> | 0.59 (0.04)<sup>a</sup> |
|  | Depressive symptoms<sup>†§</sup> | 0.00 (0.03)<sup>b-h</sup> | 0.44 (0.05)<sup>af</sup> | 0.42 (0.05)<sup>ae</sup> | 0.44 (0.05)<sup>af</sup> | 0.57 (0.05)<sup>acfgh</sup> | 0.29 (0.05)<sup>abde</sup> | 0.39 (0.05)<sup>ae</sup> | 0.4 (0.04)<sup>ae</sup> |
|  | Sleep-related impairments<sup>†</sup> | -0.06 (0.05)<sup>b-h</sup> | 0.27 (0.05)<sup>a</sup> | 0.29 (0.07)<sup>a</sup> | 0.26 (0.06)<sup>a</sup> | 0.36 (0.06)<sup>af</sup> | 0.16 (0.06)<sup>ae</sup> | 0.28 (0.05)<sup>a</sup> | 0.36 (0.05)<sup>af</sup> |
|  | Anger<sup>†</sup> | -0.01 (0.04)<sup>b-h</sup> | 0.29 (0.05)<sup>af</sup> | 0.26 (0.06)<sup>af</sup> | 0.24 (0.05)<sup>af</sup> | 0.35 (0.05)<sup>afg</sup> | 0.09 (0.05)<sup>bcdeh</sup> | 0.18 (0.05)<sup>ae</sup> | 0.26 (0.05)<sup>af</sup> |
|  | Global health<sup>‡</sup> | -0.02 (0.04)<sup>cdegh</sup> | -0.24 (0.05)<sup>dh</sup> | -0.36 (0.06)<sup>a</sup> | -0.40 (0.05)<sup>abf</sup> | -0.36 (0.05)<sup>a</sup> | -0.24 (0.05)<sup>dh</sup> | -0.32 (0.05)<sup>a</sup> | -0.36 (0.04)<sup>abf</sup> |
|  | Peer relations<sup>‡</sup> | 0.02 (0.05)<sup>bdeg</sup> | -0.29 (0.05)<sup>ac-fh</sup> | -0.12 (0.07)<sup>b</sup> | -0.11 (0.05)<sup>abf</sup> | -0.15 (0.06)<sup>abf</sup> | 0.02 (0.05)<sup>bdeg</sup> | -0.17 (0.05)<sup>af</sup> | -0.07 (0.05)<sup>b</sup> |
| DREAMS | N | - | 260-276 | 493-515 | 305-320 | 258-274 | 216-224 | 181-191 | 153-168 |
|  | Anxiety<sup>†</sup> | - | 0.86 (0.07)<sup>c-h</sup> | 1.1 (0.06)<sup>be</sup> | 1.12 (0.07)<sup>be</sup> | 1.29 (0.07)<sup>bcdg</sup> | 1.16 (0.08)<sup>b</sup> | 1.05 (0.08)<sup>be</sup> | 1.17 (0.09)<sup>b</sup> |
|  | Depressive symptoms<sup>†</sup> | - | 0.77 (0.07)<sup>c-h</sup> | 0.98 (0.06)<sup>b</sup> | 1.06 (0.07)<sup>b</sup> | 1.06 (0.07)<sup>b</sup> | 1.13 (0.08)<sup>b</sup> | 0.98 (0.09)<sup>b</sup> | 1.06 (0.09)<sup>b</sup> |
|  | Sleep-related impairments<sup>†</sup> | - | 0.64 (0.07)<sup>c-h</sup> | 0.81 (0.06)<sup>b</sup> | 0.82 (0.07)<sup>b</sup> | 0.87 (0.07)<sup>b</sup> | 0.92 (0.08)<sup>b</sup> | 0.84 (0.08)<sup>b</sup> | 0.86 (0.09)<sup>b</sup> |
|  | Anger<sup>†</sup> | - | 0.64 (0.06)<sup>c-g</sup> | 0.82 (0.06)<sup>b</sup> | 0.87 (0.06)<sup>b</sup> | 0.87 (0.06)<sup>b</sup> | 0.89 (0.07)<sup>b</sup> | 0.82 (0.08)<sup>b</sup> | 0.79 (0.08) |
|  | Global health<sup>‡</sup> | - | -0.65 (0.06)<sup>d-h</sup> | -0.76 (0.05)<sup>defh</sup> | -0.97 (0.06)<sup>bc</sup> | -0.91 (0.06)<sup>bc</sup> | -0.94 (0.07)<sup>bc</sup> | -0.86 (0.07)<sup>b</sup> | -1.00 (0.08)<sup>bc</sup> |
|  | Peer relations<sup>‡</sup> | - | -0.42 (0.07) | -0.35 (0.06)<sup>dh</sup> | -0.53 (0.06)<sup>c</sup> | -0.45 (0.07) | -0.5 (0.07) | -0.46 (0.08) | -0.58 (0.08)<sup>c</sup> |

Note. <sup>a,b,c,d,e,f,g,h</sup> represent significant differences at $p < .05$ between measurements using Least Significant Differences post-hoc tests. E.g., superscript <sup>b</sup> in column (d) indicates a significant post-hoc difference between columns (b) and (d) for a variable.
\*Sample sizes vary because data from different domains comes from different norm studies.
<sup>†</sup> Higher scores indicate more symptoms
<sup>‡</sup> Higher scores indicate better functioning
<sup>§</sup> = Analyses are not controlled for birth country of parents and educational level parents due to missing data.

In the main text, we report the overall ANCOVA covering all time points, interactions across all time points, and comparisons between post-pandemic and pre-pandemic measurements (first pandemic measurement for DREAMS, which has no pre-pandemic data). For completeness, the Tables and Figures represent data from all time points.

## Results

### Parent-reported outcomes (BPM)

Table 2 presents the results for the BPM outcome measures of the NTR and DREAMS samples, and Figure 2 illustrates the EMMs of the general population sample and clinical sample over time, represented as standard deviations from pre-pandemic NTR norm scores.

**Figure 2.**
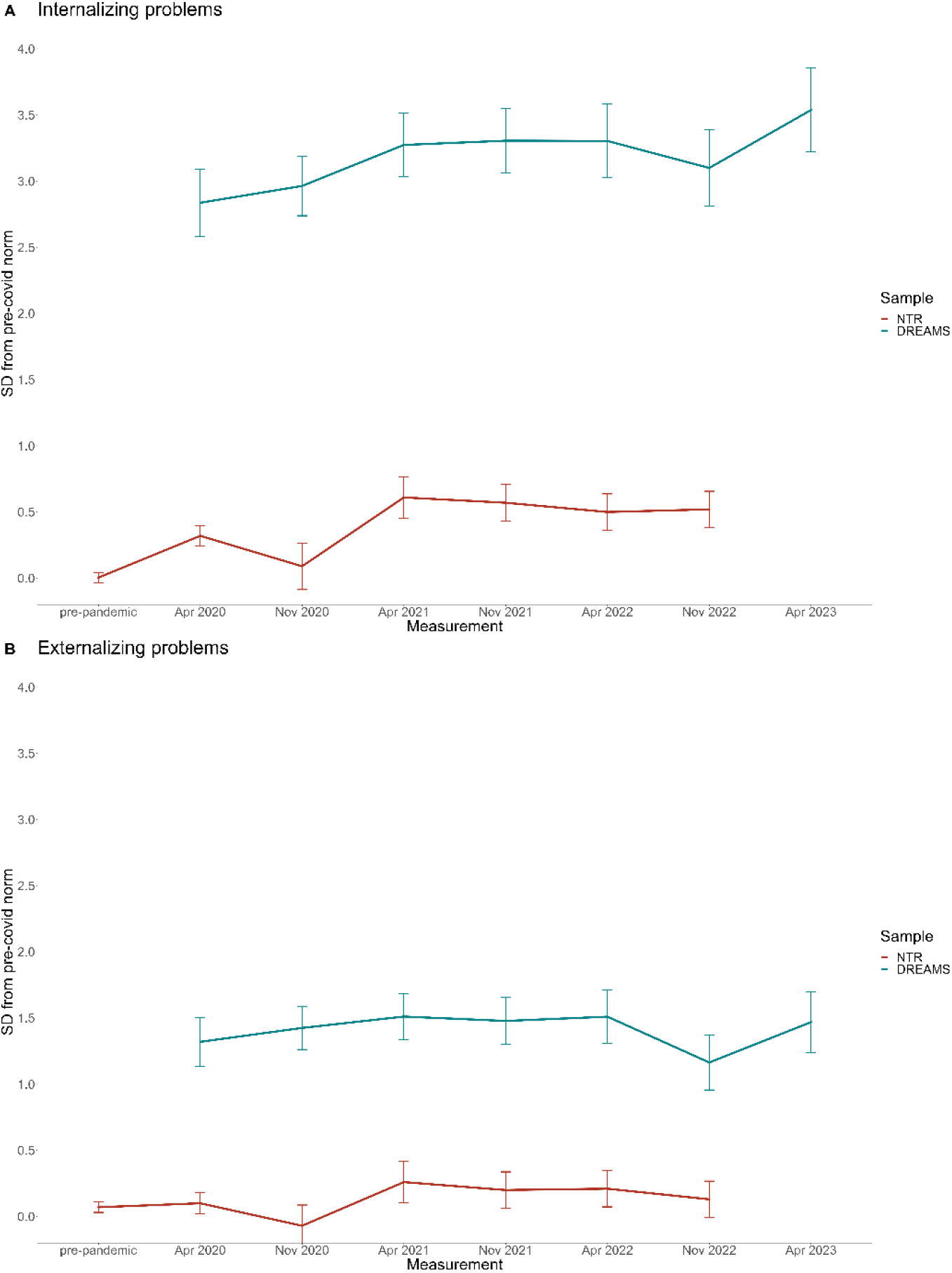

In the general population sample of NTR, internalizing problems differed significantly between measurements (*p* < .001). There was a significant interaction between time and age (*p* < .05), where younger children’s problems varied less between measurements compared to older children. There was no interaction between time and sex. On the post-pandemic measurement of NTR (Nov/Dec 2022), scores were significantly higher than during the pre-pandemic measurement (*p* < .001).

Externalizing problems also differed significantly between measurements (*p* < .01). We found no interactions between time and age nor time and sex. On the post-pandemic measurement scores did not differ significantly from the pre-pandemic measurement.

In the clinical sample of DREAMS (note that there were no pre-pandemic data available for this sample), internalizing problems differed significantly between measurements (*p* < .001). We found no interactions between time and age nor time and sex. On the first post-pandemic measurement, scores did not differ significantly from the first COVID measurement. In the second post-pandemic measurement of DREAMS, scores were significantly higher than during the first COVID measurement (*p* < .001).

Externalizing problems did not significantly differ between measurements, and we found no interactions between time and age nor time and sex.

### Child-reported outcomes (PROMIS)

Table 2 presents the results for the PROMIS outcome measures of the KLIK and DREAMS samples, and Figure 3 illustrates the EMMs of the general population sample and clinical sample over time, represented as standard deviations from pre-pandemic norm scores. Table S2 shows the EMMs of the U.S. calibrated T-scores for international comparison.

**Figure 3.**
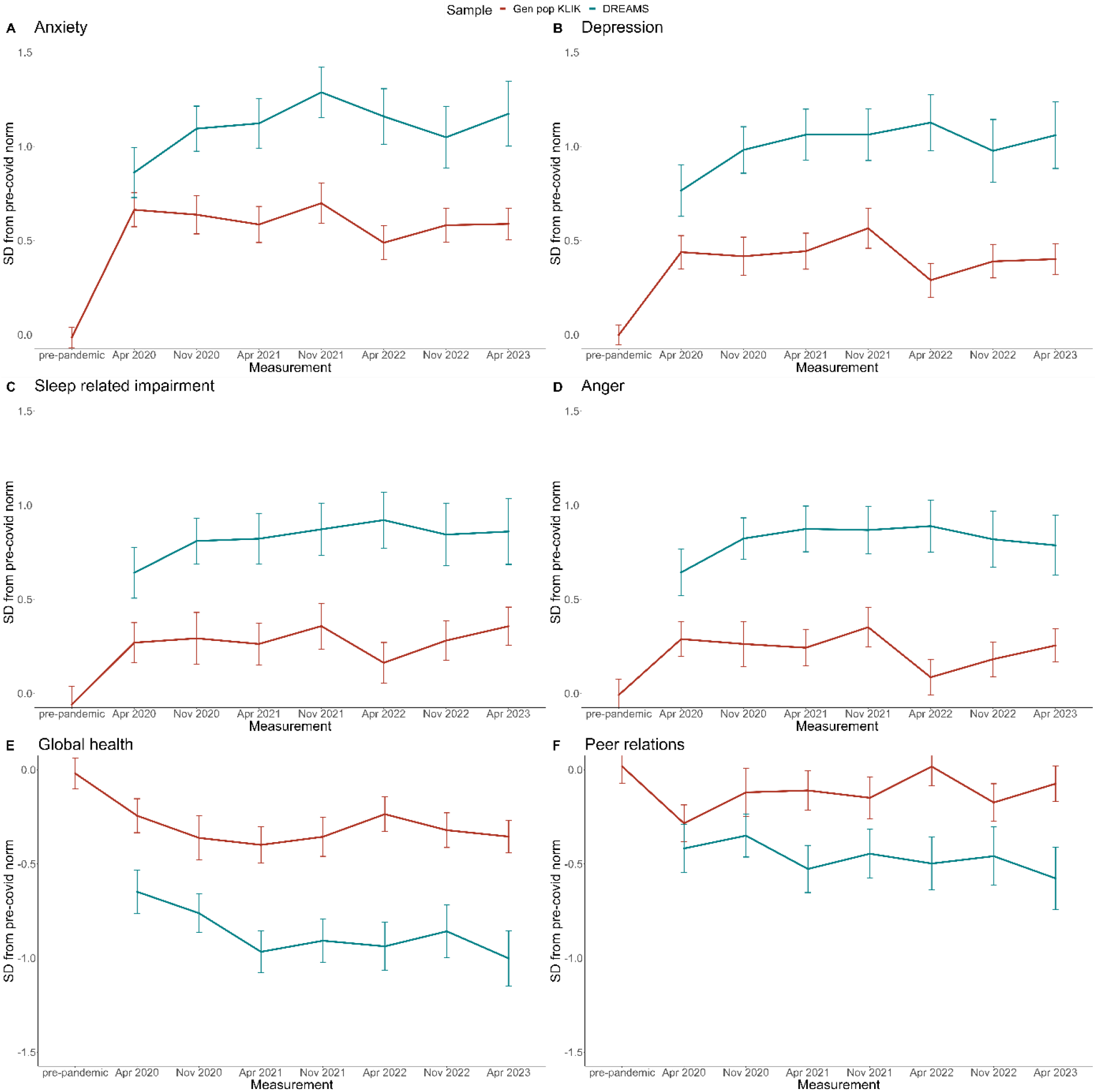

In the KLIK sample, scores on all six PROMIS domains differed significantly between measurements (all *p*s < .001). We found significant interactions (all *p*s < .01) between time and age for Sleep-related impairment (*p* < .01), where younger children showed a larger increase in impairment from the pre-pandemic measurement to the first COVID measurement, and for Global health (*p* < .01), where younger children showed a larger increase in impairment throughout the pandemic.

For Anxiety, Depression, Sleep-related problems, Anger, and Global health, scores on both post-pandemic measurements were significantly worse than those during the pre-pandemic measurement (*p*s < .01). For Peer relations, scores were significantly worse on the first post-pandemic measurement (*p* < .001), but not the second, compared to the pre-pandemic measurement.

In the DREAMS sample (note that there were no pre-pandemic data available for this sample), scores on all PROMIS domains except Peer relations differed significantly between measurements (all *p*s < .05). We found a significant interaction between time and age for Sleep-related impairment (*p* < .05), where older children showed a larger increase in impairment throughout the pandemic.

For Anxiety, Depression, Sleep-related problems, and Global health, scores on both post-pandemic measurements were significantly worse than those on the first pandemic measurement (all *p*s < .05). For Anger, scores were significantly worse on the first post-pandemic measurement (*p* < .05), but not the second, compared to the first pandemic measurement.

## Discussion

In this study, we investigated whether a previously observed trend during the COVID-19 pandemic towards pre-pandemic levels of mental health in the general population and increases in internalizing mental health problems in children in psychiatric care have continued. We described parent-reported and self-reported child mental health at seven or eight (depending on sample) cross-sectional measurements, ranging from pre-pandemic (2018-2019) to post-pandemic (November 2022 and April 2023) periods, in two general population samples and one clinical sample from the Netherlands (age 8 to 18 years).

We found no changes in externalizing problems as reported by parents in the general population, but parents did report higher internalizing problems post-pandemic than pre-pandemic. Children also reported increased mental health problems post-pandemic, especially in anxiety and depression, to a lesser extent in sleep-related impairment and global health, and least in anger. Thus, the previously observed trend towards pre-pandemic levels of mental health (Zijlmans et al., 2023; Deng et al., 2023) did not continue in our samples. These findings can be interpreted in several ways. First, the enduring impact of the COVID pandemic may stem from disruptions in daily routines, school progress, and social interactions, and therefore a lack of perspective, next to economic or medical challenges faced by families. Second, it is possible that the increase in mental health problems is not only specific to the pandemic but reflects a broader trend. However, longitudinal studies utilizing long-term historical data suggest that the pandemic has exacerbated these trends (Fischer et al. 2022; Kiviruusu et al. 2023). Third, increased societal attention to mental health during the pandemic might have influenced the reporting of mental health problems, indicating a rise in awareness, openness, and willingness to report mental health problems rather than due to a genuine increase in the prevalence of mental health issues.

In the clinical sample receiving psychiatric care, we also found no changes in externalizing problems in the cohorts over time. However, we did observe increased internalizing problems reported by parents throughout the pandemic, with the highest scores at the second post-pandemic measurement (April 2023). A similar pattern is present in the child self-reported data that show greatest increases in problems in anxiety, depression, and global health, to a lesser extent on sleep-related impairment, and least on anger. Similar to the general population sample, we found no improvement in the post-pandemic measurements. In addition to the possible explanations mentioned for the general population sample, in the clinical sample there are more unknown factors that might contribute to the observed trend. For instance, it is possible that the population seeking psychiatric care during the COVID-19 period had more severe problems compared to previous years, resulting in differences in populations. Moreover, the shift from in-person to online psychiatric care for many children during the pandemic and the increased demands on health care services may have impacted the efficacy of therapy. Lastly, the absence of pre-pandemic data for the clinical sample limits our ability to compare the magnitude of increases in problems relative to the pre-pandemic period.

Like our previous findings, we found no evidence for sex effects over time and minor evidence for age effects over time, but the latter is inconsistent throughout the samples and outcomes. Although it has been suggested that girls and older children may be more vulnerable to effects of the pandemic, these may simply be representative of established associations between sex, age, and mental health rather than differences occurring due to the pandemic. For example, a Finnish study on anxiety in 750,000 adolescents (aged 13-20 years) found no sex effects over time when controlling for existing trends (Kiviruusu et al. 2023).

The strengths of this study lie in the inclusion of multiple large samples from both general and clinical populations, the systematic measurement of child mental health at seven time points over the course of three years via validated self-report and parent-report measures, and the ability to compare the general population samples with pre-pandemic data. The study also has several limitations. First, there is a risk of selection bias in the NTR and DREAMS samples, as response rates were limited. Second, the use of independent cross-sectional measurements, while minimizing contamination of treatment and developmental effects, limits our ability to investigate different trajectories over time. Finally, no pre-pandemic data were available for the DREAMS sample.

Overall, the impact of the COVID-19 pandemic on child mental health is complex and requires rigorous investigation (Cortese, Solmi, and Correll 2023). While we can now conclude that the pandemic has had both acute and longer-term impact on child mental health, it remains somewhat unclear which specific aspects of the pandemic have contributed to these effects and which children were most vulnerable or resilient. Comparative studies between countries with different lockdown policies, particularly regarding school closures, are needed to disentangle putative effects. Two meta-analyses suggested school closures had a specific negative effect, but data were limited to the first year of the pandemic and other factors were not tested (Ludwig-Walz et al. 2022, 2023). Longitudinal studies are needed to assess distinct trajectories and differentiate between children who are more severely impacted and those who may have even benefited. Finally, consistent monitoring of child mental health remains crucial so that policies on (national and international) interventions, education, and clinical care can be guided by empirical data.

## Conclusion

Child mental health problems in the general population are substantially higher post-pandemic compared to pre-pandemic measurements and the previously observed trend towards pre-pandemic levels has not continued. In children in psychiatric care mental health problems have increased during the pandemic and are substantially higher post-pandemic than at the start of the pandemic. Longitudinal and comparative studies are needed to assess what the most important drivers of these changes are. Monitoring of child mental health is important to identify vulnerable groups more easily and to be able to respond to acute demands more quickly.

## Acknowledgements

We thank all participating families. This research was funded by ZonMw project number 50-56300-98-973. Data collection by KLIK (measurement 1 and 2) was supported by Stichting Steun Emma Kinderziekenhuis. PROMIS reference data collection was supported by the National Health Care Institute and the Netherlands Organization for Health Research and Development. Data collection in the NTR was supported by: NWO large investment (480-15-001/674; Netherlands Twin Registry Repository: researching the interplay between genome and environment). MB is supported by a European Research Council consolidator Grant (WELL-BEING 771057 PI Bartels).

## DATA AVAILABILITY STATEMENT

All data produced in the present study are available upon reasonable request to the authors.

## CONFLICTS OF INTEREST STATEMENT

MB is supported by a European Research Council consolidator Grant (WELL-BEING 771057 PI Bartels). The authors have declared that they have no competing or potential conflicts of interest.

All authors have seen and approved the manuscript.

