## Supplemental Tables for "Changes in child and adolescent mental health across the COVID-19 pandemic (2018-2023): Insights from general population and clinical samples in the Netherlands"

*Table S1. BPM parent-report sum score estimated marginal means (EMM), standard errors, comparisons between measurement points, and % elevated scores*

| Cohort | | 0 (a) pre-pandemic | 1 (b) Apr 2020 | 2 (c) Nov 2020 | 3 (d) Apr 2021 | 4 (e) Nov 2021 | 5 (f) Apr 2022 | 6 (g) Nov 2022 | 7 (h) Apr 2023 |
|---|---|---|---|---|---|---|---|---|---|
| NTR | N | 13345 | 1296 | 220 | 297 | 372 | 376 | 329 | - |
| | BPM Internalizing | 0.88 (0.03)[bd-g] | 1.35 (0.06)[ac-g] | 1.01 (0.13)[bd-g] | 1.79 (0.12)[abc] | 1.73 (0.1)[abc] | 1.63 (0.1)[abc] | 1.66 (0.11)[abc] | - |
| | BPM Externalizing | 2.16 (0.04)[df] | 2.21 (0.08) | 1.88 (0.17)[def] | 2.54 (0.16)[ac] | 2.42 (0.13)[c] | 2.45 (0.14)[ac] | 2.29 (0.14) | - |
| | BPM Int % elevated | 7.0% | 15.4% | 12.3% | 21.2% | 22.8% | 18.9% | 17.9% | - |
| | BPM Ext % elevated | 8.6% | 8.8% | 5.9% | 12.1% | 10.3% | 10.5% | 9.1% | - |
| DREAMS | N | - | 431 | 675 | 538 | 488 | 355 | 338 | 294 |
| | BPM Internalizing | - | 5.12 (0.2)[defh] | 5.31 (0.17)[defh] | 5.77 (0.18)[bc] | 5.81 (0.19)[bc] | 5.81 (0.21)[bc] | 5.51 (0.22)[h] | 6.16 (0.24)[bcg] |
| | BPM Externalizing | - | 4.7 (0.19) | 4.92 (0.17)[g] | 5.09 (0.18)[g] | 5.02 (0.18)[g] | 5.09 (0.21)[g] | 4.39 (0.22)[c-fh] | 5.01 (0.24)[g] |
| | BPM Int % elevated | - | 62.4% | 64.0% | 72.7% | 72.3% | 71.6% | 72.5% | 72.5% |
| | BPM Ext % elevated | - | 38.1% | 39.1% | 39.6% | 41.2% | 40.9% | 35.5% | 39.8% |

Note. [a,b,c,d,e,f,g,h] represent significant differences at $p < .05$ between measurements using Least Significant Differences post-hoc tests. E.g., superscript [b] in column (d) indicates a significant post-hoc difference between columns (b) and (d) for a variable.

*Table S2. PROMIS self-report T-score estimated marginal means (EMM), standard errors, and comparisons between measurement points*

| Cohort | | 0 (a) pre-pandemic | 1 (b) Apr 2020 | 2 (c) Nov 2020 | 3 (d) Apr 2021 | 4 (e) Nov 2021 | 5 (f) Apr 2022 | 6 (g) Nov 2022 | 7 (h) Apr 2023 |
|---|---|---|---|---|---|---|---|---|---|
| KLIK | N | 527-1082* | 467-482 | 408-423 | 383-387 | 335-345 | 448-455 | 474-485 | 551-577 |
| | Anxiety† | 43.7 (0.3)[b-h] | 50.2 (0.4)[af] | 50 (0.5)[af] | 49.5 (0.5)[a] | 50.6 (0.5)[af] | 48.5 (0.5)[abce] | 49.5 (0.4)[a] | 49.5 (0.4)[a] |
| | Depressive symptoms† | 44.7 (0.3)[b-h] | 49.4 (0.5)[af] | 49.1 (0.6)[ae] | 49.4 (0.5)[af] | 50.7 (0.6)[acfgh] | 47.8 (0.5)[abde] | 48.8 (0.5)[ae] | 49 (0.4)[ae] |
| | Sleep-related impairments† | 47 (0.5)[b-h] | 50.3 (0.5)[a] | 50.5 (0.7)[a] | 50.2 (0.6)[a] | 51.2 (0.6)[af] | 49.2 (0.6)[aeh] | 50.4 (0.5)[a] | 51.2 (0.5)[af] |
| | Anger† | 44.1 (0.5)[b-h] | 47.5 (0.5)[af] | 47.2 (0.7)[af] | 47 (0.6)[af] | 48.2 (0.6)[afg] | 45.2 (0.5)[bcdeh] | 46.3 (0.5)[ae] | 47.1 (0.5)[af] |
| | Global health‡ | 46.4 (0.4)[cdegh] | 45.9 (0.5)[dh] | 44.8 (0.6)[a] | 44.4 (0.5)[abf] | 44.8 (0.5)[a] | 46 (0.5)[dh] | 45.2 (0.5)[a] | 44.8 (0.4)[abf] |
| | Peer relations‡ | 47.1 (0.4)[bdeg] | 44.2 (0.5)[ac-fh] | 45.8 (0.6)[b] | 45.9 (0.5)[abf] | 45.5 (0.5)[abf] | 47.1 (0.5)[bdeg] | 45.2 (0.5)[af] | 46.2 (0.5)[b] |
| DREAMS | N | - | 260-276 | 493-515 | 305-320 | 258-274 | 216-224 | 181-191 | 153-168 |
| | Anxiety† | - | 52.16 (0.66)[c-h] | 54.42 (0.59)[be] | 54.7 (0.65)[be] | 56.29 (0.67)[bcdg] | 55.05 (0.73)[b] | 53.98 (0.8)[be] | 55.19 (0.86)[b] |
| | Depressive symptoms† | - | 52.82 (0.74)[c-h] | 55.11 (0.66)[b] | 55.97 (0.73)[b] | 55.97 (0.74)[b] | 56.65 (0.81)[b] | 55.06 (0.9)[b] | 55.93 (0.96)[b] |
| | Sleep-related impairments† | - | 54.01 (0.69)[c-h] | 55.7 (0.62)[b] | 55.82 (0.68)[b] | 56.32 (0.7)[b] | 56.81 (0.76)[b] | 56.04 (0.84)[b] | 56.2 (0.89)[b] |
| | Anger† | - | 51.53 (0.72)[c-g] | 53.58 (0.64)[b] | 54.16 (0.7)[b] | 54.09 (0.73)[b] | 54.33 (0.79)[b] | 53.53 (0.87)[b] | 53.17 (0.93) |
| | Global health‡ | - | 41.95 (0.57)[d-h] | 40.83 (0.51)[defh] | 38.82 (0.56)[bc] | 39.41 (0.58)[bc] | 39.11 (0.63)[bc] | 39.9 (0.69)[b] | 38.48 (0.74)[bc] |
| | Peer relations‡ | - | 42.93 (0.62) | 43.57 (0.56)[dh] | 41.89 (0.61)[c] | 42.67 (0.63) | 42.17 (0.68) | 42.54 (0.75) | 41.41 (0.8)[c] |

Note. [a,b,c,d,e,f,g,h] represent significant differences at $p < .05$ between measurements using Least Significant Differences post-hoc tests. E.g., superscript [b] in column (d) indicates a significant post-hoc difference between columns (b) and (d) for a variable.

* Sample sizes vary because data from different domains comes from different norm studies.

† Higher scores indicate more symptoms

‡ Higher scores indicate better functioning

*Table S3. PROMIS % normal, moderately elevated, and severely elevated scores*

| | | KLIK | | | | | | | | DREAMS | | | | | | | |
|---|---|---|---|---|---|---|---|---|---|---|---|---|---|---|---|---|---|
| | | 0 (a) pre-pandemic | 1 (b) Apr 2020 | 2 (c) Nov 2020 | 3 (d) Apr 2021 | 4 (e) Nov 2021 | 5 (f) Apr 2022 | 6 (g) Nov 2022 | 7 (h) Apr 2023 | 0 (a) pre-pandemic | 1 (b) Apr 2020 | 2 (c) Nov 2020 | 3 (d) Apr 2021 | 4 (e) Nov 2021 | 5 (f) Apr 2022 | 6 (g) Nov 2022 | 7 (h) Apr 2023 |
| N | | 527-1082* | 467-482 | 408-423 | 383-387 | 335-345 | 448-455 | 474-485 | 551-577 | - | 260-276 | 493-515 | 305-320 | 258-274 | 216-224 | 181-191 | 153-168 |
| Anxiety | normal | 74.8% | 45.9% | 53.0% | 53.8% | 52.5% | 59.7% | 58.5% | 55.8% | - | 46.5% | 33.5% | 34.1% | 29.3% | 33.6% | 35.9% | 34.2% |
| | moderate | 20.0% | 51.2% | 42.9% | 36.6% | 35.0% | 32.7% | 33.3% | 32.9% | - | 40.1% | 43.6% | 48.4% | 45.2% | 44.5% | 48.9% | 44.9% |
| | severe | 5.2% | 2.9% | 4.1% | 9.6% | 12.5% | 7.5% | 8.1% | 11.3% | - | 13.4% | 23.0% | 17.5% | 25.6% | 21.8% | 15.2% | 20.9% |
| Depressive Symptoms | normal | 74.8% | 60.6% | 61.1% | 57.3% | 53.4% | 67.1% | 62.9% | 61.1% | - | 53.4% | 39.2% | 38.9% | 37.3% | 35.5% | 41.2% | 36.4% |
| | mod | 20.1% | 36.9% | 35.0% | 36.7% | 36.8% | 27.3% | 31.2% | 33.2% | - | 30.7% | 32.8% | 36.9% | 44.1% | 38.2% | 38.5% | 39.0% |
| | severe | 5.1% | 2.6% | 3.9% | 6.0% | 9.8% | 5.6% | 5.9% | 5.7% | - | 15.9% | 28.0% | 24.2% | 18.6% | 26.4% | 20.3% | 24.7% |
| Sleep-Related impairment | normal | 71.5% | 61.8% | 57.2% | 57.4% | 53.1% | 64.8% | 58.0% | 55.9% | - | 51.5% | 40.0% | 42.3% | 39.6% | 36.5% | 37.9% | 36.4% |
| | moderate | 23.3% | 36.0% | 39.9% | 38.1% | 40.4% | 31.8% | 35.9% | 36.9% | - | 37.4% | 44.4% | 43.9% | 45.8% | 45.7% | 50.5% | 50.6% |
| | severe | 5.1% | 2.1% | 2.9% | 4.4% | 6.5% | 3.3% | 6.1% | 7.2% | - | 11.1% | 15.6% | 13.8% | 14.6% | 17.8% | 11.5% | 13.0% |
| Anger | normal | 75.0% | 74.9% | 72.2% | 69.9% | 65.2% | 78.9% | 72.1% | 70.6% | - | 54.7% | 46.6% | 41.9% | 42.3% | 42.7% | 44.5% | 42.3% |
| | moderate | 19.9% | 21.9% | 22.7% | 24.4% | 28.4% | 17.5% | 23.7% | 22.6% | - | 30.2% | 34.4% | 40.3% | 41.6% | 40.5% | 40.7% | 41.7% |
| | severe | 5.1% | 3.2% | 5.1% | 5.7% | 6.4% | 3.5% | 4.2% | 6.8% | - | 15.1% | 19.0% | 17.7% | 16.1% | 16.8% | 14.8% | 16.0% |
| Global Health | normal | 75.0% | 74.3% | 64.3% | 66.9% | 69.3% | 73.4% | 69.9% | 69.2% | - | 60.9% | 45.4% | 41.3% | 44.5% | 38.8% | 40.8% | 38.1% |
| | moderate | 20.1% | 23.7% | 29.1% | 28.9% | 24.3% | 21.5% | 24.1% | 24.8% | - | 27.2% | 29.7% | 34.4% | 33.2% | 32.6% | 33.5% | 40.5% |
| | severe | 5.0% | 2.1% | 6.6% | 4.1% | 6.4% | 5.1% | 6.0% | 6.1% | - | 12.0% | 24.9% | 24.4% | 22.3% | 28.6% | 25.7% | 21.4% |
| Peer relations | normal | 74.2% | 70.7% | 80.6% | 77.8% | 71.6% | 82.6% | 73.6% | 76.2% | - | 65.8% | 67.5% | 57.4% | 61.6% | 60.6% | 56.9% | 54.2% |
| | moderate | 19.9% | 24.6% | 14.0% | 17.2% | 21.2% | 12.7% | 17.9% | 18.3% | - | 21.9% | 23.5% | 32.1% | 29.8% | 30.6% | 32.0% | 34.0% |
| | severe | 5.9% | 4.7% | 5.4% | 5.0% | 7.2% | 4.7% | 8.4% | 5.4% | | 12.3% | 8.9% | 10.5% | 8.5% | 8.8% | 11.0% | 11.8% |

* Sample sizes vary because data from different domains comes from different norm studies.
